# Duration of referral-to-death and its influencing factors among cancer and non-cancer patients: perspective from a community palliative care setting in Malaysia

**DOI:** 10.1101/2022.08.17.22277787

**Authors:** Yan Yee Yip, Wen Yea Hwong, Sylvia Ann McCarthy, Aidah Abdul Hassan Chin, Yuan Liang Woon

## Abstract

**Objectives:** To compare the referral-to-death duration among palliative cancer and non-cancer patients and to determine its influencing factors in a Malaysian community hospice.

**Methods:** This retrospective cohort study included decedents referred to palliative care in a community hospice between January 2017 to December 2019. Referral-to-death is the interval between first referral date to date of death. Besides descriptive analyses, negative binomial regression analyses were conducted to identify factors associated with referral-to-death duration among both groups.

**Results:** Of 4346 patients referred, 86.7% (n=3766) and 13.3% (n=580) had primary diagnoses of cancer and non-cancer respectively. Median referral-to-death was 32 days (IQR:12-81) among cancer patients and 19 days (IQR:7-78) among non-cancer patients. The shortest referral-to-death duration among cancer patients were for liver cancer (Median:22 days,IQR:8-58.5). Non-cancer patients with dementia, heart failure and multisystem failure had the shortest referral-to-death duration at 14 days. Among cancer patients, longer referral-to-death duration was associated with women compared to men (IRR:1.26,95%CI:1.16-1.36) and patients aged 80 to 94 years old compared to below 50 years old (IRR:1.19,95%CI:1.02-1.38). Cancer patients with analgesics prescribed before palliative care had 29% fewer palliative care days compared to those with no analgesics prescribed before referral. Non-cancer patients aged 50 to 64 years old had shorter referral-to-death duration compared to below 50 years old (IRR:0.51,95%CI:0.28-0.91).

**Conclusion:** Shorter referral-to-death duration among non-cancer patients indicated possible access inequities with delayed palliative care integration. Factors influencing referral-to-death duration should be accounted for in developing targeted approaches to ensure timely and equitable access to all patients requiring palliative care.

**KEY MESSAGES:** 

**What is already known on this topic:**

⍰ Addressing the need for timely integration of palliative care in low- and middle-income countries (LMIC) is a priority, considering its increasing burden of non-communicable diseases with limited data available in this region.

**What this study adds:**

⍰ Findings from our study have shown an underrepresentation of non-cancer patients amongst community palliative care referrals with a shorter referral-to-death duration among these patients as compared to cancer patients.
⍰ Age, sex, and use of analgesia prior to referrals were factors significantly associated with referral-to-death duration among cancer patients whereas for non-cancer patients, older aged patients had a shorter referral-to-death duration..

**How this study might affect research, practice or policy:**

⍰ The underrepresentation of non-cancer referrals for palliative care indicates the need to determine reasons for the disparity in referrals between cancer and non-cancer patients in clinical practice and to evaluate feasible and effective approaches to narrow this gap in settings similar to that of ours.
⍰ At a policy level, plans to develop interventions to allow timely integration of community palliative care should target specific groups of patients.

## INTRODUCTION

Palliative care is a multidisciplinary approach that provides specialized medical and nursing care for patients with life-threatening illnesses. Palliative care enhances patients’ and carers’ quality of life, and alleviates symptom burden in patients with an active and progressive disease with low cure rates.[1] Among adults, some of the chronic diseases in need of palliative care include cardiovascular diseases (38.5%) and cancer (34%).[2] Palliative Care has been included in universal health care by World Health Assembly (WHA) since 2014 and as such is a human right. In recent years, global palliative care needs have inflated due to aging of the world population and an increase in global disease burden.[3] Global cancer burden for example, is projected to be 28.4 million cases in 2040.[4]

Due to the growing importance of palliative care, equitable access is crucial for all patient groups across high-income and low- and middle-income countries (LMIC). Overall, 40 million people are estimated to be in need of palliative care yearly, with 78% coming from LMIC as a result of having a huge burden of non-communicable diseases (NCD) deaths (31.4 million).[5] Nevertheless, scarce resources in this region has led to challenges and dilemma in the prioritization of services among different patient groups. Access to palliative care services in LMIC has been reported to be limited, covering only 14% of those in need.[2]

Among patients with access privileges, referral to palliative care should be personalized around their needs, and delivered at a timely integration at a suitable place.[6] An early and timely integration of palliative care would reduce unnecessary hospital admissions, medical treatments and financial costs.[7] Evidence from randomized trials has suggested that continuity of palliative care with a multidisciplinary team for 3 to 4 months is required to elicit full benefits.[8] Nevertheless in real world practice, only about 50% of patients were referred less than 19 days before death, with a large variation between countries, from 6 days in Australia to 69 days in Canada.[7] Besides a known disparity between guidelines and clinical practice, duration of palliative care may also vary due to inclusion of patients with varied disease progression. NICE review guidelines have indicated that referral period for metastatic cancer patients could be range from only a few weeks to the last 6 to 12 months of life.[9]

In Malaysia, palliative care development is at a stage of preliminary integration into mainstream service provision.[10] Public-private partnership is viewed as a long term aim in providing seamless end-of-life patient care.[11] This is especially so as Malaysia is poised to become an aging nation over the next decade,[12] with palliative care needs projected to escalate from 71,675 patients in 2004 to 144,454 in 2019, and to approximately 239,713 patients in 2030.[13] In 1992, palliative care in Malaysia was started by non-governmental organizations (NGO), providing community palliative care.[14] Besides a lack of equity and standardization, they operate as individual entities, catering to patients in urban areas. In 2016, government-based domiciliary palliative care (DPC) programmes began in 4 states, under the initiative of the Ministry of Health. Issues with workforce and funding has meant that development of these services has been slow.[15] Despite facing numerous challenges, community-based palliative care is preferred by patients where these services are available, as majority prefer to be cared for at home.[16]

There are limited studies within the LMIC region examining timely integration of community palliative care from the aspects of referral-to-death intervals.[7] In Malaysia, there are no previous studies looking at the differences between cancer and non-cancer patients receiving community palliative care. Comparison of palliative care timelines between these patient groups allows a better understanding of their differences and provides evidence-based information to address gaps in palliative care services within the country as well as countries of similar settings.

Therefore, in this study, we compared the duration of referral-to-death among cancer and non-cancer patients and determined factors influencing the duration length in the largest community palliative care centre in Malaysia.

## METHODS

### Data source

Retrospective cohort data for all patients referred to Hospis Malaysia for community palliative care from 1st of January 2017 to 31st December 2019 were extracted from the electronic medical record system by the hospice information technologist on 19th August 2020. This marked the last available information for patients dated 18th August 2020.

### Patient selection

Patients with first referral date within the study period who had passed away at time of reporting (18th August 2020) were included in the study. Patients who were still receiving care at time of reporting were excluded.

Of 7005 patients, 4766 patients had died at time of reporting. 4346 patients were eligible for main analysis as 420 patients did not have primary diagnosis stated. Of those, 4306 (99.1%) patients were selected for palliative timeline analysis and a further 4277 (98.4%) patients with no missing socio-demographics were included in subsequent regression analysis. Flow chart of patient recruitment is outlined in Appendix 1.

### Determinants and outcome

We extracted the following information from the system: 1) patients’ characteristics upon referral (age, gender, primary diagnosis, referral location), 2) prescription of analgesics prior to first referral, 3) place of death, and 4) palliative care timelines comprising of date of first referral and date of death.

The referral-to-death duration was used as a proxy to study the main outcome which is timing of palliative care referral. This interval was defined as duration between the first palliative care referral date to date of death. Exact appropriate timing for referral has always been difficult to establish but several studies have measured this interval as an indicator to assess the appropriateness of referral timelines for palliative care.[17-18] For the analysis of this outcome, 40 patients had negative referral-to-death values and were excluded. This was most likely a representation of patients who have passed away before their scheduled first referral dates.

### Statistical analysis

Data analysis was conducted using descriptive statistics. Categorical variables were expressed in frequency and percentages; while numerical data was presented as median with interquartile range (IQR) for non-normally distributed variables.

Pearson Chi-Square (χ2) test compared patient’s characteristics between cancer and non-cancer patients. Because 8% of patients in the cohort were excluded due to not having any primary diagnosis, we compared the characteristics of those included and excluded in the main analysis due to missingness (Appendix 2). Secondly, referral-to-death duration was estimated in median and IQR stratified by patient’s characteristics and diagnoses. Comparison between cancer and non-cancer patients was performed using Mann-Whitney test. Thirdly, multivariable negative binomial regression analyses were conducted to identify factors which were associated with referral-to-death duration among cancer and non-cancer patients. A negative binomial model was chosen to adjust for the overdispersed count outcome data.

All statistical testing were 2-sided and a p-value<0.05 was considered statistically significant. RStudio® *Version 1*.*4*.*1564* was used for data analysis.

## RESULTS

### Baseline characteristics

Table 1 illustrates the baseline characteristics of patients referred for palliative care. Of 4346 patients, 86.7% were patients with cancer. Lung cancer, colorectal cancer and breast cancer were the top three diagnoses. The remaining 13.3% patients had non-cancer as primary diagnosis, of these 35.9% were patients with renal failure. (Appendix 3) Patients with non-cancer were older with 81.1% aged over 65 years, compared to 56.3% patients with cancer. In both cancer and non-cancer patients, the proportion of male and female patients were similar, with slightly more female patients being referred for palliative care services. Among cancer patients, government hospital referrals topped the referral category (38.6%) whereas for non-cancer patients, the highest referral came from university hospital (46.4%). More than half of cancer and non-cancer patients died at home, at 56.8% and 61.6% respectively. Less than one-tenth of cancer patients (7.1%) were prescribed with analgesics prior to their first referral compared to 5.2% of non-cancer patients.

**Table 1:**
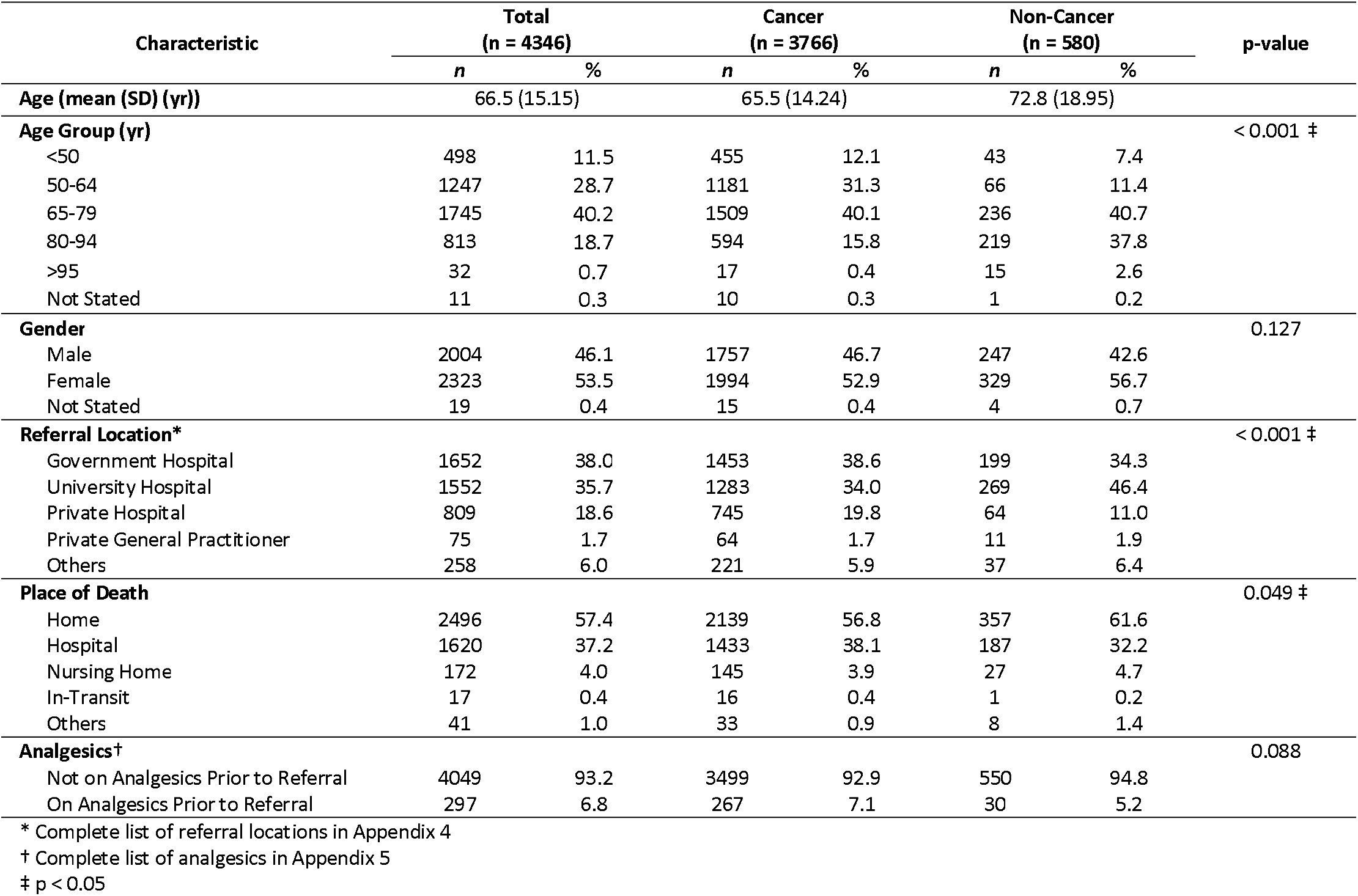
Baseline characteristics of palliative care cancer and non-cancer patients (n = 4346)

### Median duration between referral-to-death

Table 2 depicts the median referral-to-death duration for cancer and non-cancer patients, further stratified by patient’s characteristics. Median duration was 32 days (IQR:12-81) for cancer patients and significantly shorter among non-cancer patients at 19 days (IQR:7-78).

**Table 2:**
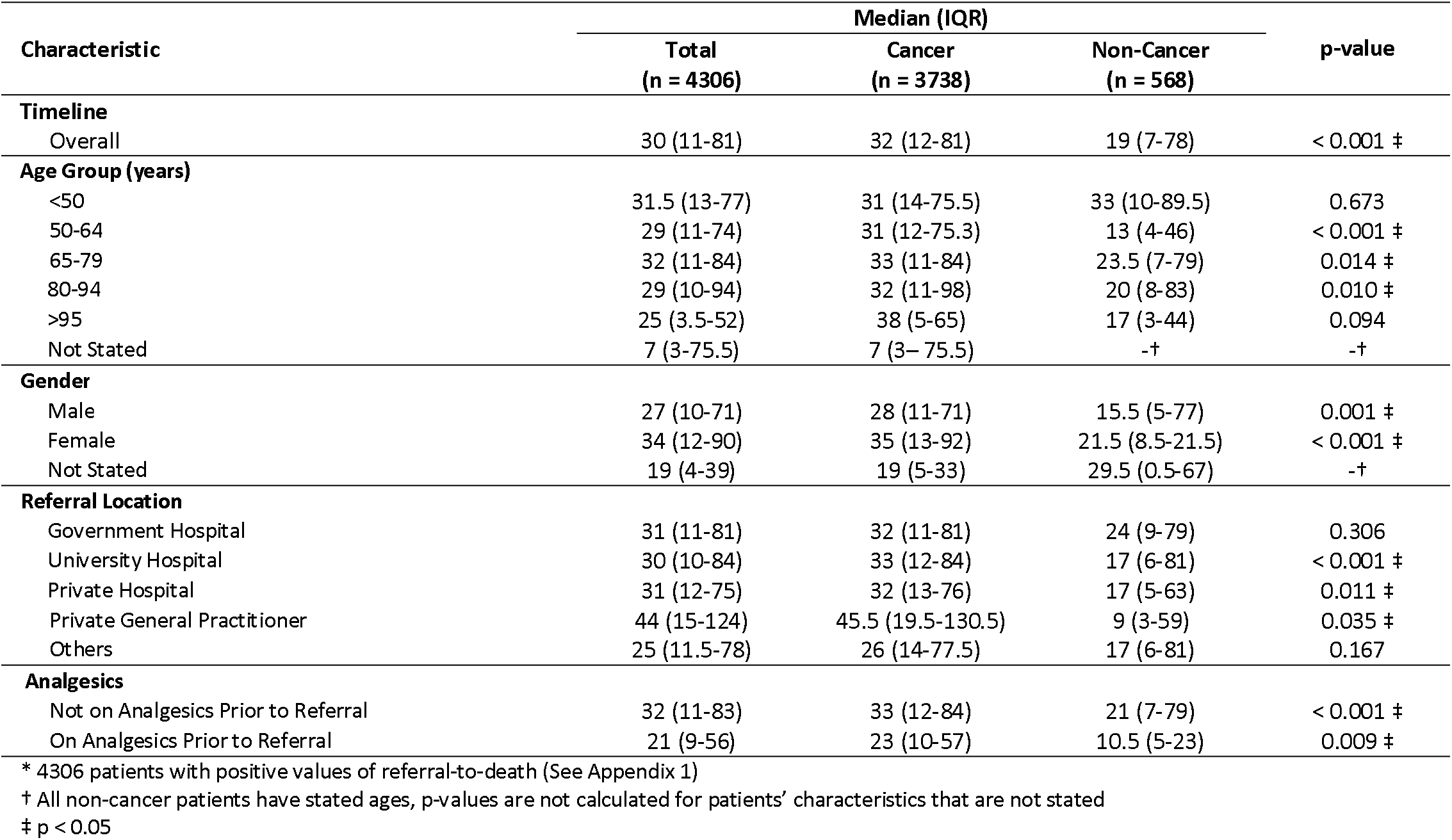
Median duration between referral-to-death for cancer and non-cancer patients (n = 4306)*

The 1-month duration was similar for cancer patients of all age groups and for non-cancer patients aged less than 50 years. There was a significant difference in predefined age groups, where the duration was significantly shorter in non-cancer patients compared to cancer patients; in the 50 to 64 age group median duration was 31 days (IQR:12-75.3) for cancer patients and 13 days (IQR:4-46) for non-cancer patients (p<0.001), 65 to 79 years (33 days (IQR:11-84) in cancer versus 23.5 days (IQR:7-79) in non-cancer), and 80 to 94 years (32 days (IQR:11-98) in cancer versus 20 days (IQR:8-83) in non-cancer).

Women had a longer median referral-to-death duration, 34 days (IQR:12-90) compared to men with 27 days (IQR:10-71). Similar patterns were observed in each group. Male patients with cancer had significantly longer referral-to-death duration of 12.5 days compared to men with non-cancer diagnosis.. Similarly, the duration was longer by 13.5 days among female patients with cancer than women with non-cancer diagnosis. (p<0.001).

There were significant differences between both patient groups referred from university hospitals, private hospitals and private general practitioners (GPs). The largest difference was noted among private GPs referrals, taking into consideration the wide confidence intervals due to the smaller number of patients in the group. Cancer patients showed longer median referral-to-death of 45.5 days (IQR:19.5-130.5) than the overall timeline, while non-cancer patients had significantly shorter median referral-to-death duration at 9 days (IQR:3-59).

Of those on analgesics prior to referral, median duration between referral-to-death was 23 days (IQR:10-57) among cancer patients and 10.5 days (IQR:5-23) among non-cancer patients (p=0.009).

### Patients’ referral-to-death stratified by primary diagnoses

Figure 1 shows that the shortest referral-to-death duration among cancer patients was for liver cancer (22 days (IQR:8-58.5)), followed by pancreatic cancer (23.5 days (IQR:10.5-52.5)) and haematological cancer (25 days (IQR:7-69)). Brain cancer and head and neck cancer had the longest median referral-to-death duration at 50 days (IQR:20.5-124) and 45 days (IQR:14-100.5), respectively.

**Figure 1.**
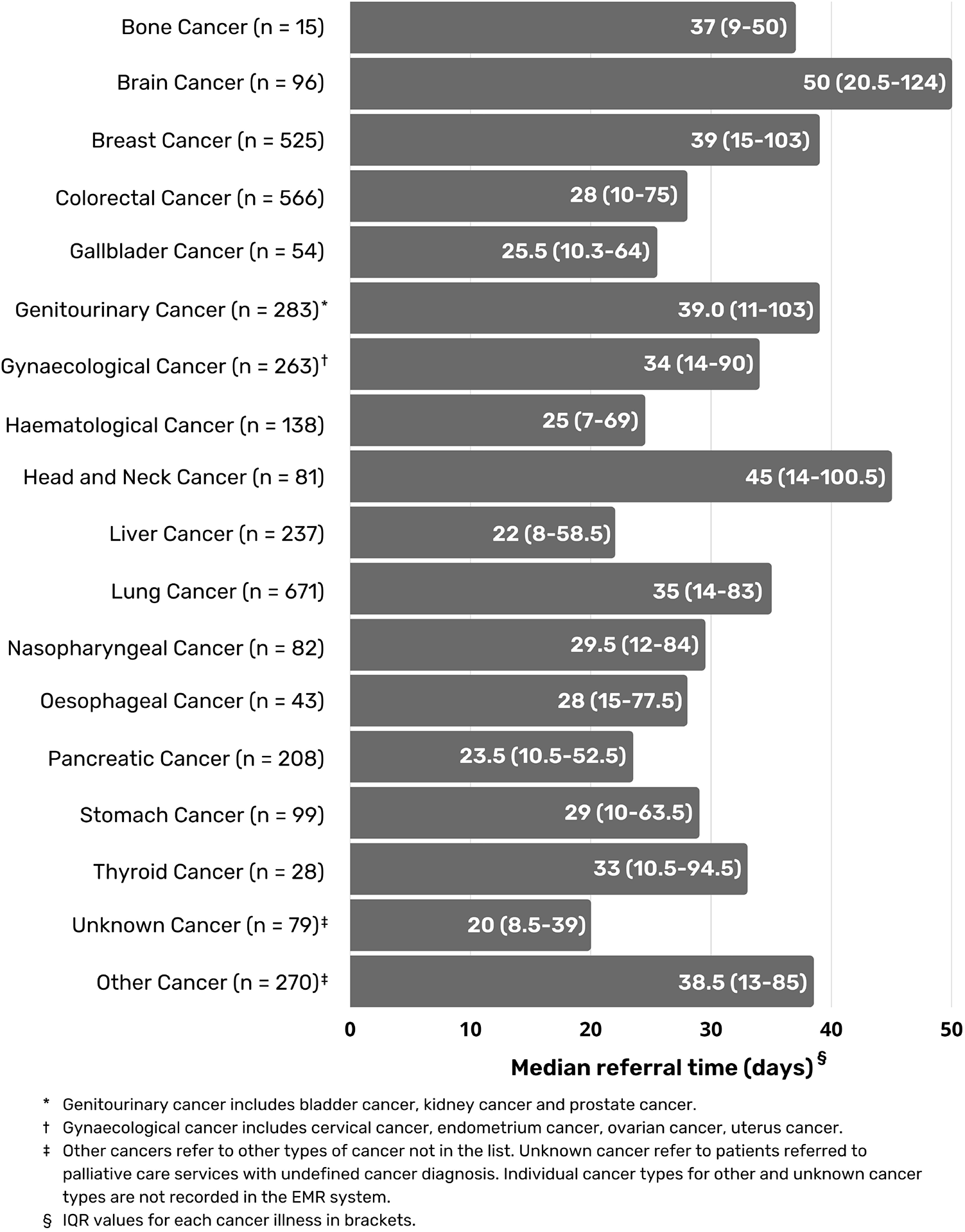
Median referral-to-death stratified by individual cancer diagnosis (n = 3738).

As for non-cancer, patients with dementia, heart failure and multisystem failure had the shortest referral-to-death duration at 14 days (Figure 2). Only 3 non-cancer diagnoses had median referral-to-death intervals of more than 30 days, with the longest duration among patients with chronic obstructive pulmonary disorder at 101 days (IQR:20-290).

**Figure 2.**
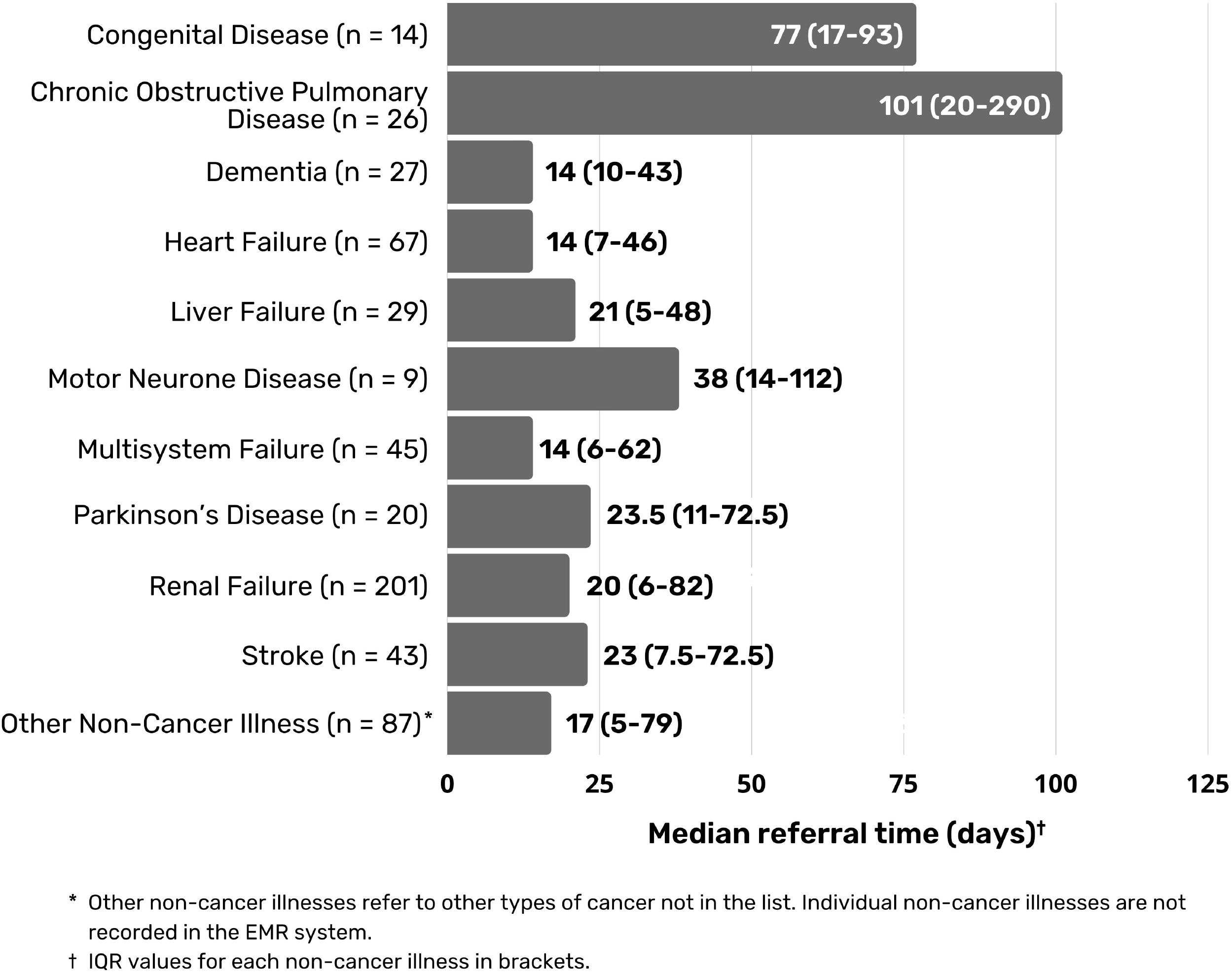
Median referral-to-death stratified by individual non-cancer diagnosis (n = 568).

### Factors influencing referral-to-death

Multivariable negative binomial models were used to examine the association between patient variables and referral to-death duration for cancer and non-cancer patients (Table 3). Among cancer patients, patients aged 80 to 94 years old had 19% more days of hospice-based palliative care compared to those below 50 years old (IRR:1.19,95%CI:1.02-1.38). In contrast, other age groups showed shorter palliative care days when compared to cancer patients aged 50 years old and below, although these did not reach statistical significance. Referral-to-death duration was also 26% longer for females compared to males (IRR:1.26,95%CI:1.16-1.36). Among cancer patients with analgesics prescribed before palliative care referral, referral-to-death duration was 29% shorter (IRR:0.71,95%CI:0.61-0.82) as compared to those without. For non-cancer patients, older patients above the age of 50 years old had shorter referral-to-death duration compared to patients below 50 years old, with the largest difference of 49% shorter palliative care days for patients aged 50 to 64 years old (IRR:0.51,95%CI:0.28-0.91).

**Table 3:**
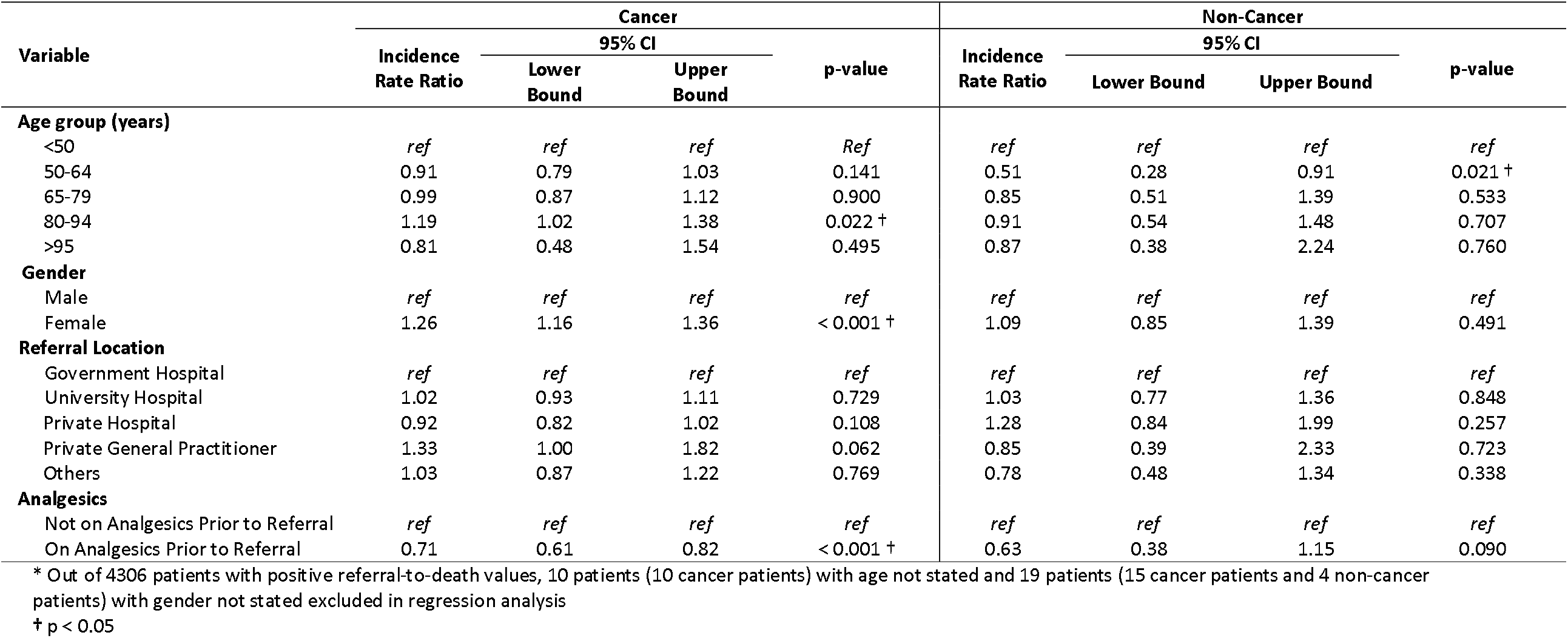
Negative binomial analyses of referral-to-death (n = 4277)*

## DISCUSSION

To the best of our knowledge, this is the first study in Malaysia where an assessment of timely referrals to palliative care among cancer and non-cancer patients in a community palliative care was conducted. Our findings showed an underrepresentation of non-cancer amongst community palliative care referrals, with only one-tenth of referred patients having non-cancer diagnoses. We found that the referral-to-death duration was one month for cancer patients, with a shorter duration of approximately two weeks among non-cancer patients. Among cancer patients, older age between 80 to 94 years, females and patients who were prescribed with analgesia prior to first referral had longer durations of referral-to-death. Contrastingly, older age among non-cancer patients was associated with shorter duration of palliative care days.

Our study has shown that cancer diagnosis is the primary determinant of palliative care access. Consistent with a report published by London School of Economics and Political Science for the year 2012 to 2013, cancer patients were more frequently referred to palliative care services in England, Wales and Northern Ireland than non-cancer patients, despite cancer accounting for only 29% of deaths in the region.[19] Similarly, despite having a comparable number of patients dying from non-cancer illnesses to patients dying from cancer in Australia, unmet palliative care access was mostly seen in the former.[20] As for the perspectives of LMIC, there is limited information on palliative care access for non-cancer patients. Overall, the underrepresentation of non-cancer patients for referrals to palliative care services were largely attributed to barriers to access which include lack of clear referral criteria guidelines for non-cancer patients, inability to identify non-cancer patients who can be referred to palliative care due to unexpected disease progression with different pathways of care,[21] and inability to accommodate non-cancer patients due to lack of resources (e.g. lack of non-cancer expertise among palliative care staff).[22] Parallel to what we found, non-cancer patients were often only referred at an older age, when their prognosis are poorer, leading to a shorter referral-to-death.[22] This demonstrates the fact that despite a general understanding of the extension of palliative care services beyond cancer care, it has yet to be commonly exercised by physicians.[23] This is a potential missed opportunity to optimise the quality of life for non-cancer patients.

We found an overall duration between referral to death of 32 days but to date, there is no consistency in the definition of a timely duration for palliative care or the method of its assessment to allow possible benchmarking for improvement of quality of care. We adapted a commonly used assessment of this duration where earlier access to palliative care is associated with improved patient quality of life (QoL).[8] A marker of patient QoL includes determining palliative care duration, which informs access to palliative care and its potential benefits from the services.[24] Longer referral-to-death duration has been linked to better end of life outcomes.[24]

A systematic review of randomized trials focusing on early integration of palliative care has suggested that the optimal provision of palliative care services, universally favoured in healthcare policy to reach maximum patient benefits, has to be at least 3 to 4 months before death.[8] However, the duration depicted in randomized trials did not reflect the actual situation in real-world studies, with a systematic review of 169 observational studies encompassing 24 countries reporting a median of 20 days for community palliative care.[7] Our findings were consistent with the duration reported from countries with similar palliative care development (Stage 4a using the WHPCA palliative care development) at a median of 28 days.[7] Similarly, a study in Taiwan which is also a Stage 4a country has shown that the median duration to hospice palliative care was at 28 days.[25]

One notable finding from our study was the shorter referral-to-death at 19 days among non-cancer patients in comparison to cancer patients. This finding is consistent with a study conducted in United Kingdom hospices which showed a significantly shorter referral-to-death interval among non-cancer than cancer patients (27 days vs 53 days, p<0.0001).[26] Similarly, a retrospective study in the United States reported a significantly shorter median enrolment-to-death interval between patients with end-stage heart failure as compared to those with cancer (12 days vs 20 days, p<0.001).[27] All these findings reflect a tendency for late referrals among non-cancer patients, despite evidence of commonalities of palliative care needs and symptom burden between both groups.[28]

Addressing factors influencing the length of referral-to-death duration would enable us to identify predictors for early referral to allow palliative care to meet its aspirations in providing universal benefit. Based on previous studies, we would expect an increasing age being a significant predictor of shorter palliative care duration.[26, 29] A study in Leeds, United Kingdom found that older people are more disadvantaged in access to care which leads to shorter palliative care duration.[29] While we reported consistent findings among non-cancer patients, our results showed an unexpected association of longer palliative care days among cancer patients between 80 and 94 years of age as compared to those below 50 years of age. Establishing a cohesive reason for such differences remains a challenge. We postulated the possibility of having multiple chronic debilitating diseases as a reason for earlier referrals for this group of patients. Dementia for example, is prominent in one-quarter of patients aged 85 and above.[30]

Furthermore, female cancer patients had a significantly longer referral-to-death duration than their male counterparts. Reasons for this are possibly related to either the types of cancer referrals or health-seeking behaviours. As observed in our study findings, the referral-to death duration for women-related cancers which include breast and gynaecological cancers were observed to be longer than the average palliative care timeline for cancer patients. Previous studies have also highlighted the higher likelihood of women to be enrolled for hospice or palliative care.[31-32] This has been attributed to the societal norms that seeking palliative care is perceived as an act of losing courage and men should be seen as tough and do not seek help.[33] Besides, women were found to be more proactive in decision making during communication with oncologists which provided them better understanding in their illness to agree to early palliative care.[34]

The role of analgesics includes acute and chronic pain relief. Opioids are also used in the management of dyspnoea. As analgesia plays an integral role in palliative care, patients who require pain and symptom management should be referred early to palliative care to allow better management of symptoms.[35] Nevertheless, our findings showed that patients on analgesics prior to palliative care referral had a shorter referral-to-death duration. We postulated that use of analgesics prior to referral suggests a more advanced disease severity at the point of referral. Besides, they may have received care and analgesic prescribing from other services such as hospital-based palliative care and are referred to community palliative care only when their function deteriorates and are unable to attend hospital appointments.

We believe that this is the first study in the country which focuses on timely palliative care referral in a community hospice setting. Although our study data was sourced from one centre, this is the largest community hospice in Malaysia with the highest number of registered palliative care patients.[36] Therefore, our findings should provide a general picture of referrals to community palliative care in the country. Nevertheless, caution should also be taken as generalisability is subjected to variation in locality, quality, practices and standards of care in different institutions. One other limitation from this study is the inability to account for factors which are potentially important to better understand the referral-to-death duration but are not available or have huge amounts of missingness such as ethnicity, patient comorbidities and socio-economic statuses.

Findings from our study have several implications related to research, policy and clinical practice. In line with the 2019-2030 National Care Policy and Strategy Planning,[37] first, there is a crucial need to determine reasons for the underrepresentation of non-cancer patients in palliative care referrals in our local setting. A study in the United Kingdom has identified reasons for limited referrals among non-cancer patients to range from system level barriers including lack of resources and restrictive eligibility criteria to individual factors such as healthcare providers’ perception of palliative care services. Depending on the similarity or differences in these barriers, feasible and effective approaches to allow for timely introduction of palliative care, in particular among non-cancer patients, should be developed. Furthermore, these interventions should also be target-specific, accounting for the factors that influenced early referral such as age, gender, and use of analgesia. Second, it is important to target approaches to educate and improve awareness on the benefits of early palliative care amongst healthcare providers, patients and carers. Thirdly, the element of research is crucial to be included to ensure that the proposed interventions or approaches are adequately evaluated on their effectiveness and feasibility prior to their implementation.

## CONCLUSION

In conclusion, the shorter referral-to-death duration among non-cancer patients compared to cancer patients indicated a possible inequity of access with delayed integration of palliative care. Age, gender and use of analgesia before referrals were associated with this duration. These factors should be accounted for in developing targeted approaches to ensure a timely and equitable access to all patients requiring palliative care.

## Data Availability

All data produced in the present study are available upon reasonable request to the authors.

## ACKNOWLEDGMENTS

The authors would like to thank the Director General of the Ministry of Health Malaysia for his permission to publish this article.

## FOOTNOTES

### Contributors

YYY, WYH and YLW have contributed to the conception or design of the study; YYY and AAHC have contributed to data collection; YYY and WYH have contributed to the data analysis; YYY, WYH, SAM, AAHC and YLW have contributed to data interpretation; YYY drafted the manuscript; YYY, WYH, SAM, AAHC and YLW were involved in critical revision of the manuscript. All authors gave their final approval of the version to be published.

## Provenance and Peer Review

Not commissioned; externally peer reviewed.

## Supplemental Material

This content has been supplied by the author(s). It has not been vetted by BMJ Publishing Group Limited (BMJ) and may not have been peer-reviewed. Any opinions or recommendations discussed are solely those of the author(s) and are not endorsed by BMJ. BMJ disclaims all liability and responsibility arising from any reliance placed on the content. Where the content includes any translated material, BMJ does not warrant the accuracy and reliability of the translations (including but not limited to local regulations, clinical guidelines, terminology, drug names and drug dosages), and is not responsible for any error and/or omissions arising from translation and adaptation or otherwise.

## DATA AVAILABILITY STATEMENT

Data are available on reasonable request. Patient data were obtained from Hospis Malaysia on request.

## ETHICS STATEMENT

### Patient Consent for Publication

Not required.

### Ethics Approval

This study was conducted in accordance with the ethical principles outlined in the Declaration of Helsinki and the Malaysian Good Clinical Practice Guideline. The study was registered under the National Medical Research Register (NMRR-19-3472-51996) and approved by the Medical Research Ethics Committee, Ministry of Health Malaysia.

