## Supplemental Material for "Duration of referral-to-death and its influencing factors among cancer and non-cancer patients: perspective from a community palliative care setting in Malaysia"

### APPENDICES

**Appendix 1:** Flowchart of patient recruitment into study

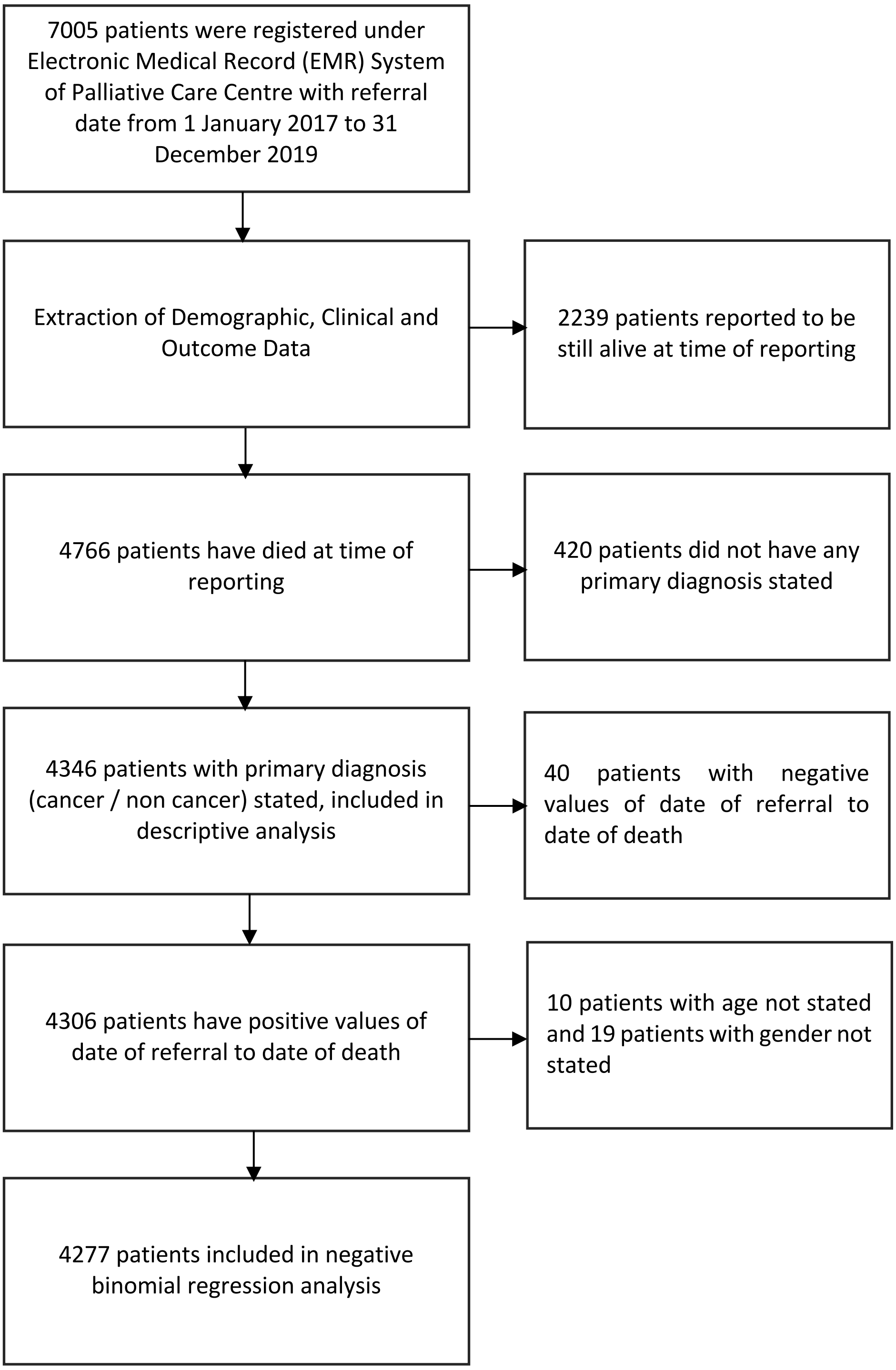

**Appendix 2:** Comparisons of patients' characteristics with primary diagnosis (n = 4346) and with no primary diagnosis (n = 420)*

| Characteristic | Patients with Primary Diagnosis (n = 4346) | | Patients with No Primary Diagnosis (n = 420) | |
|---|---|---|---|---|
| | *n* | % | *n* | % |
| **Age (mean (SD) (yr))** | 66.5 (15.15) | | 64.2 (14.69) | |
| **Age Group (yr)** | | | | |
| <50 | 498 | 11.5 | 53 | 12.6 |
| 50-64 | 1247 | 28.7 | 145 | 34.5 |
| 65-79 | 1745 | 40.2 | 165 | 39.3 |
| 80-94 | 813 | 18.7 | 54 | 12.9 |
| >95 | 32 | 0.7 | 1 | 0.2 |
| Not Stated | 11 | 0.3 | 2 | 0.5 |
| **Gender** | | | | |
| Male | 2004 | 46.1 | 219 | 52.1 |
| Female | 2323 | 53.5 | 196 | 46.7 |
| Not Stated | 19 | 0.4 | 5 | 1.2 |
| **Referral Location** | | | | |
| Government Hospital | 1652 | 38.0 | 186 | 44.3 |
| University Hospital | 1552 | 35.7 | 127 | 30.2 |
| Private Hospital | 809 | 18.6 | 69 | 16.4 |
| Private GP | 75 | 1.7 | 2 | 0.5 |
| Others | 258 | 6.0 | 36 | 8.6 |
| **Place of Death** | | | | |
| Home | 2496 | 57.4 | 130 | 30.9 |
| Hospital | 1620 | 37.2 | 276 | 65.7 |
| Nursing Home | 172 | 4.0 | 7 | 1.7 |
| In-Transit | 17 | 0.4 | 0 | 0 |
| Others | 41 | 1.0 | 7 | 1.7 |

* Patients with no primary diagnosis (n = 420) were excluded in study analysis, whereas patients with primary diagnosis (n = 4346) were included in descriptive study analysis.

**Appendix 3:** Primary diagnosis of patients (n = 4346)

| Primary Diagnosis | Cancer (n = 3766) | | Primary Diagnosis | Non-Cancer (n = 580) | |
|---|---|---|---|---|---|
| | *n* | % | | *n* | % |
| **Cancer** | | | **Non-Cancer** | | |
| Bone Cancer | 17 | 0.5 | Congenital Disease | 15 | 2.6 |
| Brain Cancer | 96 | 2.5 | Chronic Obstructive Pulmonary Disease | 27 | 4.6 |
| Breast Cancer | 527 | 14.0 | Dementia | 27 | 4.6 |
| Colorectal Cancer | 572 | 15.2 | Heart Failure | 67 | 11.6 |
| Gallbladder Cancer | 55 | 1.5 | Liver Failure | 30 | 5.2 |
| Genitourinary Cancer | 285 | 7.6 | Motor Neurone Disease | 9 | 1.6 |
| Gynaecological Cancer | 265 | 7.0 | Multisystem Failure | 45 | 7.8 |
| Haematology-Related Cancer | 139 | 3.7 | Parkinson's Disease | 20 | 3.4 |
| Head and Neck Cancer | 82 | 2.2 | Renal Failure | 208 | 35.9 |
| Liver Cancer | 238 | 6.3 | Stroke | 43 | 7.4 |
| Lung Cancer | 674 | 17.9 | Other Non-Cancer Illness | 89 | 15.3 |
| Nasopharyngeal Cancer | 82 | 2.2 | | | |
| Oesophageal Cancer | 43 | 1.1 | | | |
| Pancreatic Cancer | 209 | 5.5 | | | |
| Stomach Cancer | 100 | 2.7 | | | |
| Thyroid Cancer | 28 | 0.7 | | | |
| Unknown Cancer | 81 | 2.2 | | | |
| Other Cancer | 273 | 7.2 | | | |

**Appendix 4:** Patient's referral location for palliative care services (n = 4346)

| Patient's Referral Location | Frequency (n = 4346) (*n*) | Percentage (%) |
|---|---|---|
| **Government Hospital** | **1652** | **38.0** |
| Ampang Hospital | 147 | 3.4 |
| Kuala Lumpur Hospital | 338 | 7.8 |
| National Cancer Institute (NCI) | 865 | 19.9 |
| Selayang Hospital | 222 | 5.1 |
| Serdang Hospital | 53 | 1.2 |
| Sungai Buloh Hospital | 27 | 0.6 |
| **University Hospital** | **1552** | **35.7** |
| Universiti Malaya Medical Centre (UMMC) | 901 | 20.7 |
| Universiti Kebangsaan Malaysia Medical Centre (UKMMC) | 603 | 13.9 |
| Universiti Malaya Specialist Centre (UMSC) | 44 | 1.0 |
| Universiti Kebangsaan Malaysia Specialist Centre (UKMSC) | 4 | 0.1 |
| **Private Hospital** | **809** | **18.6** |
| Ampang Puteri Hospital | 8 | 0.2 |
| Assunta Hospital | 45 | 1.0 |
| Beacon Medical Centre | 100 | 2.3 |
| Columbia Asia Medical Centre | 3 | 0.1 |
| Damansara Specialist Hospital | 39 | 0.9 |
| Gleaneagles Intan Medical Centre | 71 | 1.6 |
| Pantai Bangsar Medical Centre | 124 | 2.9 |
| Pantai Cheras Medical Centre | 17 | 0.4 |
| Prince Court Medical Centre | 27 | 0.6 |
| Sentosa Medical Centre | 2 | 0.04 |
| Sime Darby Medical Centre | 135 | 3.1 |
| Sunway Medical Centre | 162 | 3.7 |
| Tawakal Hospital | 10 | 0.2 |
| Tung Shin Hospital | 66 | 1.5 |
| **Private General Practitioners** | **75** | **1.7** |
| **Others** | **258** | **5.9** |

### **Appendix 5:** List of analgesics

| Category | Medication | |
|---|---|---|
| | **Analgesic** | |
| **Paracetamol as Analgesic** | • Paracetamol | |
| **NSAIDS Analgesic** | • Benzydamine<br>• Celecoxib<br>• Diclofenac<br>• Etorixocib<br>• Flurbiprofen<br>• Ibuprofen | • Ketoprofen<br>• Mefenamic Acid<br>• Meloxicam<br>• Methylsalicylate<br>• Naproxen |
| **Opioids Analgesic** | • Buprenorphine<br>• Dihydrocodeine<br>• Fentanyl<br>• Methadone<br>• Morphine (*Routes*: IR, IV/IM, SC, SR) | • Naproxen<br>• Oxycodone (*Routes:* IR, SC, SR)<br>• Pethidine<br>• Tramadol |
| **Combination Agent** | • Codeine / Paracetamol<br>• Tramadol / Paracetamol<br>• Paracetamol / Orphenadrine | • Naproxen / Esomeprazole<br>• Oxycodone / Naloxone<br>• Pholcodeine / Promethazine |

IR: Intrarectal, IV: Intravenous, IM: Intramuscular, SC: Subcutaneous, SR: Subrectal

### **Appendix 6:** List of abbreviations

| | |
|---|---|
| **DPC** | Domiciliary Palliative Care |
| **GPs** | General Practitioners |
| **LMIC** | Low- and Middle- Income Country |
| **MOH** | Ministry of Health |
| **MREC** | Medical Research and Ethics Committee |
| **NCD** | Non-communicable Diseases |
| **NCI** | National Cancer Institute |
| **NGO** | Non-governmental Organization |
| **NMRR** | National Medical Research Registry |
| **QOL** | Quality of Life |
| **UKMMC** | University Kebangsaan Malaysia Medical Centre |
| **UKMSC** | University Kebangsaan Malaysia Specialist Centre |
| **UMMC** | University Malaya Medical Centre |
| **UMSC** | University Malaya Specialist Centre |
| **WHA** | World Health Assembly |
| **WHO** | World Health Organization |
| **WHPCA** | Worldwide Hospice Palliative Care Alliance |
